# Burden, Long-Term Trends, and Projections of Spinal Fractures in China in the Context of G20 Member Countries, 1990 – 2050: An Analysis of the Global Burden of Disease 2021 Study

**DOI:** 10.64898/2026.05.14.26353225

**Authors:** Shi-Jie Zeng, Jia-Lan Chen, Zhao-Feng Lin, Jia-Qi Zhang, Li-Xin Zhu

**Author notes:** Correspondence: Li-Xin Zhu, Department of Orthopedic Spine Surgery, Zhujiang Hospital of Southern Medical University, Guangzhou 510220, P.R. China.

## Abstract

Spinal fractures are an important contributor to disability worldwide, particularly in aging populations. However, comprehensive long-term comparisons between China and other major economies remain limited.

Using data from the Global Burden of Disease (GBD) 2021 study, we analyzed temporal trends in the incidence, prevalence, and years lived with disability (YLDs) of spinal fractures in China and the overall G20 from 1990 to 2021. Age-standardized rates were assessed using Joinpoint regression and age–period–cohort analysis. Future burden through 2050 was projected using autoregressive integrated moving average modeling, and decomposition analysis was performed to quantify the contributions of demographic and epidemiological factors.

Between 1990 and 2021, China experienced substantial increases in absolute burden. Incident cases increased by 52.27%, prevalent cases by 113.66%, and YLDs by 107.21%. The age-standardized prevalence rate (ASPR) and age-standardized YLD rate (ASYR) increased significantly, whereas the age-standardized incidence rate (ASIR) showed a non-significant upward trend. In contrast, the overall G20 aggregate showed increasing absolute case numbers but significantly declining age-standardized rates. Age–period–cohort and age-specific analyses indicated that older adults represented the main burden-bearing population. Projections suggested that China’s ASIR may decline by 2050, whereas prevalence and YLD burden, particularly among males, may remain relatively high compared with the overall G20 level. Decomposition analysis identified population aging as the major driver of burden growth.

China experienced a rising burden of spinal fractures over the past three decades, in contrast to declining age-standardized trends in the overall G20 aggregate. These findings highlight the substantial role of population aging in shaping spinal fracture burden and provide epidemiological evidence for prevention planning and aging-related health policy.

## 1. Introduction

Spinal fractures are a major cause of disability worldwide and contribute substantially to the burden of musculoskeletal disorders, particularly in older adults. These injuries are associated with chronic pain, functional limitation, reduced quality of life, and increased healthcare utilization. According to the Global Burden of Disease (GBD) 2019 study, the global incidence of vertebral fractures and the corresponding YLDs increased markedly between 1990 and 2019 [1]. Population-based studies from North America and Europe have further shown that spinal fractures are common in aging populations, underscoring their growing clinical and socioeconomic impact [2, 3].

China is experiencing rapid population aging and epidemiological transition, both of which are likely to influence the burden of spinal fractures. Previous GBD-based studies have reported global estimates and national trends in China [1, 4], and additional epidemiological investigations have been conducted in East Asia, Europe, and North America [5–11]. However, most of these studies have focused either on global patterns or on individual countries. Long-term comparative evidence placing China in the context of other major economies remains limited. As a result, China’s burden trajectory cannot be fully interpreted in relation to countries facing similar demographic aging and health system challenges.

Comparisons with G20 countries are particularly relevant because these countries account for a substantial share of the global population, economic output, and healthcare expenditure, and many are also undergoing pronounced demographic aging. Examining China alongside the G20 may therefore help identify differences in temporal trends, demographic patterns, and potential demands on healthcare systems related to fracture prevention, treatment, rehabilitation, and long-term care.

In addition, projections of future burden are important for health resource planning. Countries undergoing accelerated aging may face increasing demand for fracture-related services, including acute care, rehabilitation, and long-term management. Nevertheless, few studies have integrated historical trends, international comparisons, and future projections of spinal fracture burden within a single analytical framework.

Using data from the GBD 2021 study, we evaluated trends in the incidence, prevalence, and YLDs of spinal fractures in China from 1990 to 2021, compared these patterns with those in G20 countries, and projected the future burden through 2050. By situating China within a broader international context, this study aims to provide epidemiological evidence to support health system planning, targeted prevention strategies, and aging-related healthcare policy.

## 2. Methods

### 2.1. Data Source

Data for this study were obtained from the Global Burden of Disease (GBD) 2021 database through the Global Health Data Exchange (GHDx) platform. We extracted country-level estimates of incidence, prevalence, and years lived with disability (YLDs) for spinal fractures in China and all G20 members from 1990 to 2021. Data were stratified by sex (both sexes, male, and female), age group (all ages and 5-year age groups from <5 years to ≥95 years), and metric type (absolute numbers and age-standardized rates). Because China is included in the pooled G20 aggregate, comparisons should be interpreted as descriptive at the group level rather than fully independent between-group contrasts.

The GBD 2021 study synthesizes 100,983 data sources worldwide, including vital registration systems, verbal autopsy data, censuses, surveys, disease registries, and healthcare utilization records, and applies standardized analytical methods to estimate disease burden across locations and time. In the present study, spinal fractures were identified according to the disease category definition used in GBD 2021[12].Spinal fractures were identified according to the disease category definition used in GBD 2021. The database integrates multiple sources, including hospital records, population surveys, and literature, and does not provide detailed information on diagnostic criteria for individual fracture types. For osteoporotic fractures, cases may be identified based on radiographic findings or clinical presentation, which could lead to substantial variation in reported incidence and prevalence across locations and age groups.

For the comparative analysis, China was compared with the overall G20 aggregate, which included China. The pooled G20 aggregate was calculated using the 19 sovereign G20 member countries only. The European Union was not included in the pooled aggregate to avoid double counting of EU member states that are also individual G20 members, such as France, Germany, and Italy. Absolute numbers for the pooled G20 group were calculated by summing the estimates across all included members, whereas pooled age-standardized rates were calculated on the basis of the corresponding aggregated population structure and age-specific estimates. Because the pooled G20 estimates included China, the observed differences between China and the G20 aggregate should be interpreted as comparisons with the overall group level rather than as independent between-group contrasts.

Because this study was based exclusively on publicly available, de-identified data, ethics committee approval and informed consent were not required. This study was conducted in accordance with the Guidelines for Accurate and Transparent Health Estimates Reporting (GATHER).

### 2.2. Statistical Analysis

Age-standardized rates (ASRs), including age-standardized incidence rate (ASIR), age-standardized prevalence rate (ASPR), and age-standardized years lived with disability rate (ASYR), were used to assess spinal fracture burden. Joinpoint regression was applied to detect inflection points and segmented temporal trends in ASRs during 1990–2021. The segmented log-linear model was constructed as: ln(ASR) = α + β×t + ε, where t denotes the calendar year, ASR is the ASR in year t, α and β represent the intercept and slope of each segmented trend, respectively, and ε is the random error term. For each segment, the annual percentage change (APC) was calculated as: APC = 100×[exp(β)−1]. The average annual percentage change (AAPC) was defined as the weighted average of APCs across all segments (with the duration of each segment as the weight): AAPC = (Σ(APC×w))/Σw, where w is the number of years in each segment. The optimal number of joinpoints was selected based on the Bayesian Information Criterion (BIC). A trend was defined as upward when both the AAPC and its 95% confidence interval (CI) were > 0, downward when both were < 0, and stable otherwise [13, 14].

### 2.3. Joinpoint Regression Analysis

Joinpoint regression analysis was performed using the Joinpoint Regression Program (version 4.9.1.0; National Cancer Institute, Bethesda, MD, USA) to evaluate temporal changes in the age-standardized burden of spinal fractures. ASIR, ASPR, and ASYR were log-transformed and modeled with calendar year as the independent variable. This method identifies time points at which significant changes in linear trends occur and estimates the annual percentage change (APC) for each segment. In addition, the average annual percentage change (AAPC) and its 95% CI were calculated to summarize the overall temporal trend during the study period. APCs and AAPCs were derived by exponentiating the regression coefficients from the log-linear models, as described previously [15–18]. A trend was considered statistically significant when the corresponding 95% CI did not include 0. The number and location of joinpoints were determined using the model selection procedure implemented in the Joinpoint software.

### 2.4. Age-Period-Cohort (APC) Model

Age–period–cohort (APC) analysis was performed to evaluate the independent effects of age, period, and birth cohort on spinal fracture burden. In this framework, age effects reflect variations associated with aging and age-related physiological changes, period effects represent temporal influences affecting all age groups simultaneously, and cohort effects capture differences in risk across birth cohorts. The APC model was fitted using a Poisson log-linear model. To address the inherent identification problem caused by the exact linear dependency among age, period, and cohort, the intrinsic estimator (IE) method was applied [19, 20]. Age, period, and cohort were categorized into consecutive 5-year intervals, and relative risks (RRs) with 95% CIs were estimated. All APC analyses were conducted using Stata/MP 17.0 (StataCorp LLC, College Station, TX, USA).

### 2.5. AutoRegressive Integrated Moving Average (ARIMA) Forecasting

An autoregressive integrated moving average (ARIMA) model was applied to forecast age-standardized trends in spinal fracture burden, including ASIR, ASPR, and ASYR, from 2022 to 2050, following previous studies [21, 22]. Forecasting analyses were conducted separately by sex. Stationarity was assessed and achieved by differencing when required. Model parameters (p, d, q) were selected using the auto.arima() function in R software (version 4.2.2) [23, 24], with the Akaike information criterion (AIC) used for model selection. Model performance was assessed by residual diagnostics, including Q–Q plots, autocorrelation function (ACF) and partial autocorrelation function (PACF) plots, and the Ljung–Box test. The analysis was conducted using the forecast, tseries, and ggplot2 packages in R.

### 2.6. Decomposition Analysis

Decomposition analysis was performed to quantify the contributions of population growth, population aging, and changes in age-specific rates to temporal changes in spinal fracture burden, following previously reported approaches [25, 26]. Specifically, the changes in incident cases, prevalent cases, and YLDs between 1990 and 2021 were partitioned into three components attributable to population size, age structure, and epidemiological change. Counterfactual scenarios were constructed by sequentially replacing the population size, age structure, and age-specific rates in the baseline year with those in the comparison year, and the average contribution of each component was estimated. The results were expressed as both absolute and relative contributions of the three drivers. Visualizations were generated using the ggplot2 package in R software.

## 3. Results

### 3.1. The Overall Trends in Incidence, Prevalence, and YLDs of Spinal Fractures in China and G20 countries

As demonstrated in Table 1, the 2021 epidemiological data for spinal fractures in China (with 95% UI) were as follows: 1.1945 million incident cases (0.8890–1.5941 million), 0.7171 million prevalent cases (0.6191–0.8353 million), and 74.1 thousand YLDs (49.8–104.3 thousand). The corresponding age-standardized rates were: ASIR 76.38 per 100 000 population (95% UI: 57.33–102.58), ASPR 40.51 per 100 000 population (95% UI: 34.42–47.56), and ASYR 4.19 per 100 000 population (95% UI: 2.78–5.96).

**Table 1.**
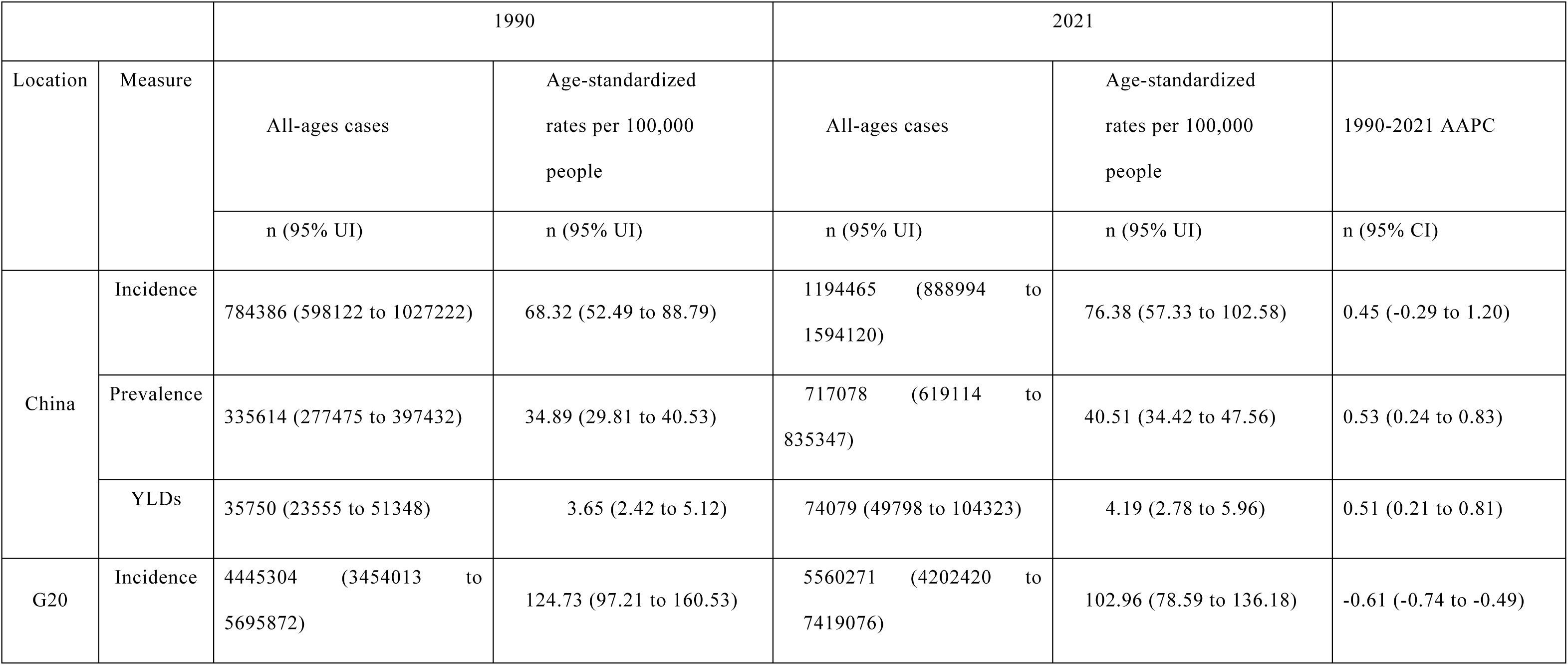
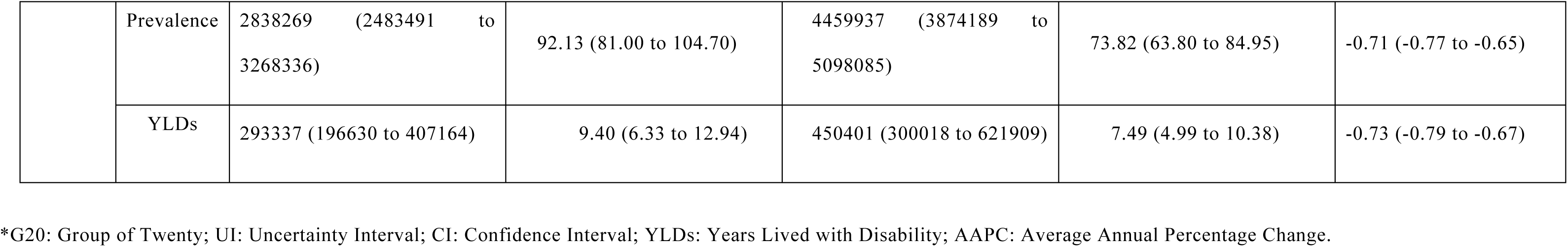
Total Age-Specific Case Numbers, Age-Standardized Incidence Rates, Prevalence Rates, YLD Rates, and Corresponding AAPCs of Spinal Fractures in China and G20 countries in 1990 and 2021.

Between 1990 and 2021, all measured indicators of spinal fractures in China exhibited varying magnitudes of increase (Table 1). All-age incident cases rose by 0.4101 million (relative increase: 52.27%), accompanied by an increase in ASIR of 8.06 per 100 000 population, with an AAPC of 0.45 (95% CI: −0.29 to 1.20), which was not statistically significant. All-age prevalent cases increased by 0.3815 million (relative increase: 113.66%), with an ASPR increase of 5.62 per 100 000 population and an AAPC of 0.53 (95% CI: 0.24 to 0.83), which was statistically significant. YLDs rose by 38.3 thousand person-years (relative increase: 107.21%), with an ASYR increase of 0.54 per 100 000 population and an AAPC of 0.51 (95% CI: 0.21 to 0.81), which was statistically significant.

For the overall G20 aggregate from 1990 to 2021, the three core indicators of spinal fractures displayed a consistent pattern: absolute case numbers increased while age-standardized rates decreased, with all reductions achieving statistical significance (Table 1). All-age incident cases increased by 1.1150 million (relative increase: 25.08%), with a corresponding decrease in ASIR of 21.77 per 100 000 population and an AAPC of −0.61 (95% CI: −0.74 to −0.49). All-age prevalent cases rose by 1.6217 million (relative increase: 57.13%), with an ASPR decrease of 18.31 per 100 000 population and an AAPC of −0.71 (95% CI: −0.77 to −0.65). YLDs increased by 157.1 thousand person-years (relative increase: 53.54%), with an ASYR decrease of 1.91 per 100 000 population and an AAPC of −0.73 (95% CI: −0.79 to −0.67).

### 3.2. Joinpoint Regression Analysis of Spinal Fracture Burden in China and G20 countries

Figure 1 presents joinpoint regression results for ASIR, ASPR, and ASYR of spinal fractures in China and G20 countries (1990–2021). China’s three age-standardized indicators showed significant fluctuations with clear joinpoints: ASIR (3 joinpoints: 2001, 2004, 2011), ASPR (4 joinpoints: 1995, 2000, 2005, 2010), and ASYR (4 joinpoints: 1995, 2000, 2005, 2011). Specifically, 2000–2005 saw statistically significant decreases in ASPR (APC = -3.59%) and ASYR (APC = -3.68%). 2010–2011 was a key turning point: 2011–2021 ASIR shifted to significant growth (APC = 2.80%), while ASPR (2010–2021: APC = 2.56%) and ASYR (2011–2021: APC = 2.59%) also increased significantly. Overall, 2011–2021 saw synchronous growth of the three indicators in China (average annual growth 2.56%–2.80%), with 1990–2021 overall AAPCs of 0.35%, 0.49%, and 0.47%, respectively.

**Figure 1.**
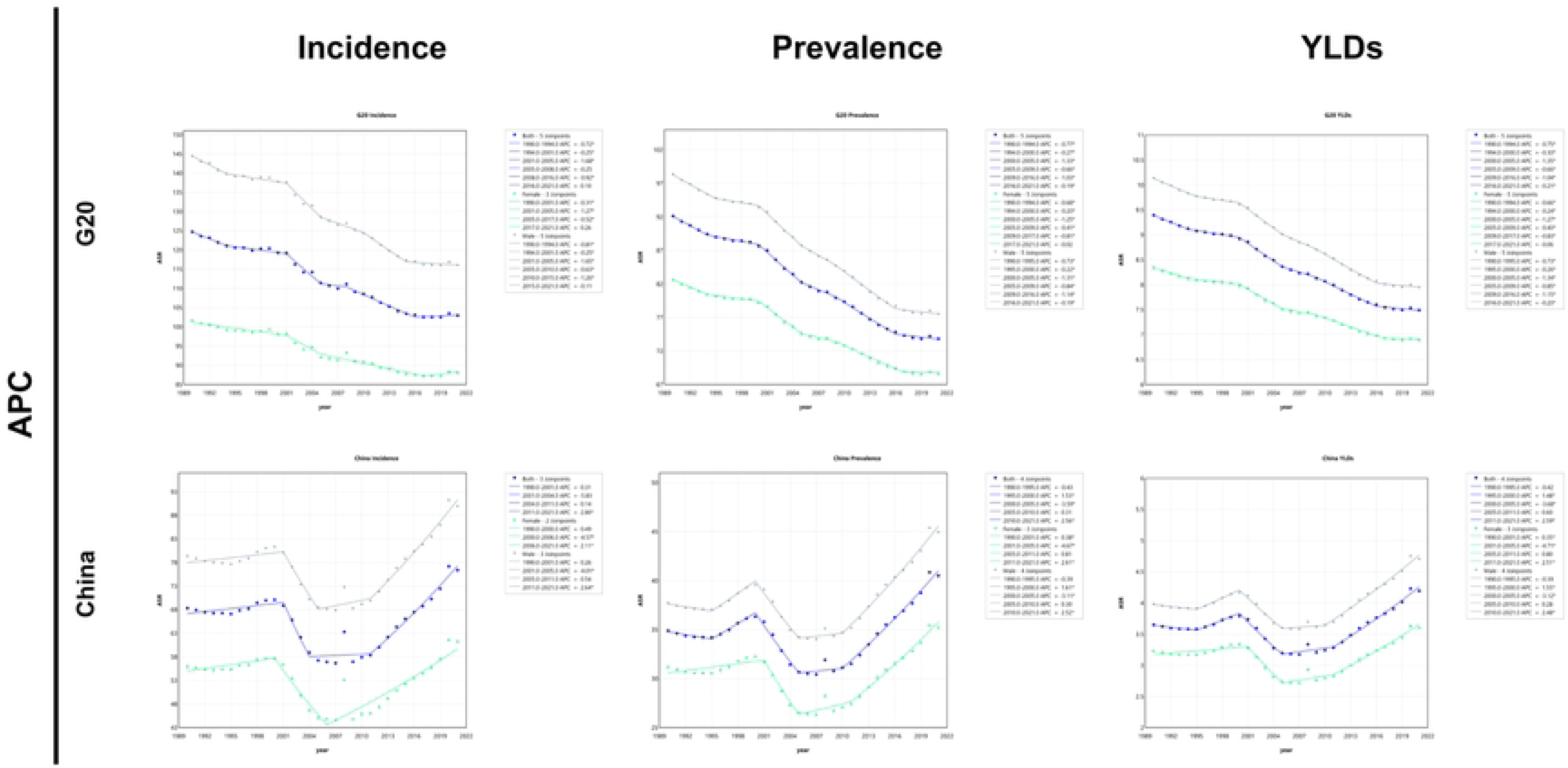
presents the Joinpoint regression analyses of ASIR, ASPR, and ASYR of spinal fractures in G20 countries and China from 1990 to 2021: (A) Joinpoint regression analysis of ASIR for spinal fractures in G20 countries; (B) Joinpoint regression analysis of ASPR for spinal fractures in G20 countries; (C) Joinpoint regression analysis of ASYR for spinal fractures in G20 countries; (D) Joinpoint regression analysis of ASIR for spinal fractures in China; (E) Joinpoint regression analysis of ASPR for spinal fractures in China; (F) Joinpoint regression analysis of ASYR for spinal fractures in China (blue=both sexes, green=females, gray=males; * indicates statistically significant APC (95% CI not including 0, P < 0.05)).

In contrast, the overall G20 aggregate showed distinct trends. 1990–2021 ASIR had a phased fluctuating downward trend (overall AAPC = -0.61%) with 5 joinpoints (1994, 2001, 2005, 2008, 2016), and a non-significant slight increase during 2016–2021 (APC = 0.10%). ASPR and ASYR maintained stable, significant downward trends throughout the study period (ASPR: overall AAPC = -0.71%; ASYR: overall AAPC = -0.73%), each with 5 joinpoints (1994, 2000, 2005, 2009, 2016). Only the annual decline rate showed phased changes (e.g., greater decline in 1990–1994, 2000–2005, 2009–2016; moderate decline in 1994–2000, 2005–2009, 2016–2021) without reversing the overall downward trend (Figure 1).

### 3.3. Comparison of Spinal Fracture Burden by Age Group and Gender between China and G20 countries

#### 3.3.1. Core Age Groups Bearing the Burden of Incident Cases

In 1990, incident cases of spinal fractures in both China and G20 countries were concentrated in the 15– 39-year age group (Figure 2 and Figure 7). By 2021, the focus of incident cases in China had shifted to the 30–59 years age group, while those in G20 countries were generally clustered in the 20–74 years age group (Figure 7). By gender, the high-incidence age group for Chinese males had migrated to 25–59 years, and the main burden interval for males in G20 countries was 15–59 years. For females, the high-risk age group for spinal fracture incidents was 30–84 years in China, compared with 50–85 years in G20 countries (Figure 2).

**Figure 2.**
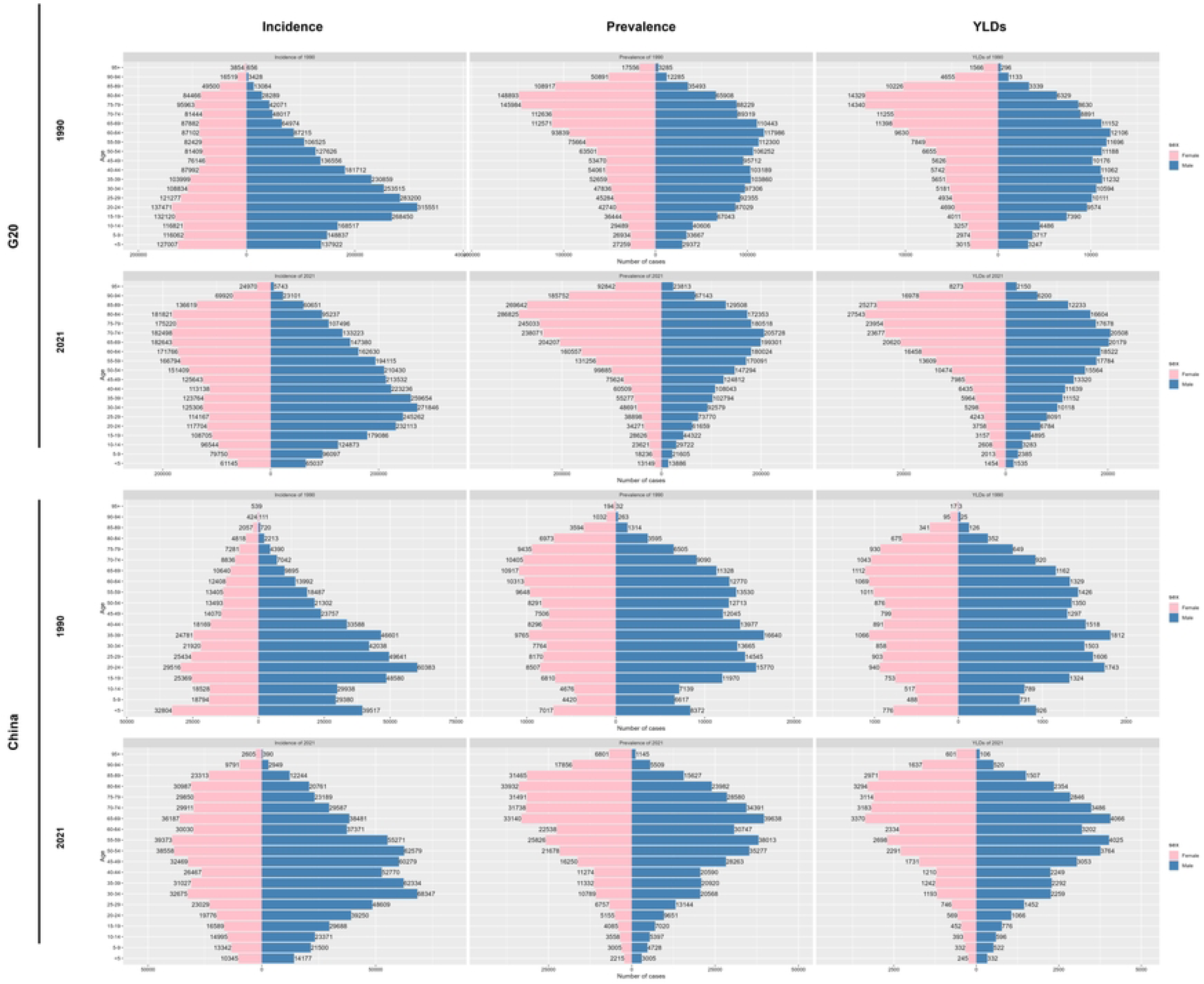
Age-sex pyramid distribution of incident cases, prevalent cases and YLDs of spinal fractures in China and G20 countries in 1990 and 2021. (A) Age-sex pyramid distribution of incident cases, prevalent cases and YLDs of spinal fractures in G20 countries in 1990 and 2021 (red for females, blue for males). (B) Age-sex pyramid distribution of incident cases, prevalent cases and YLDs of spinal fractures in China in 1990 and 2021 (red for females, blue for males).

#### 3.3.2. Burden Distribution of Prevalent Cases and YLDs

In 1990, the number of prevalent cases and YLD counts of spinal fractures among Chinese males was concentrated in the 15–69 years age group, and among females in the 55–79 years age group. By 2021, the high-burden intervals for both genders had shifted to older age groups: 40–84 years for males and 50–89 years for females (Figure 2). In contrast, throughout the study period, the high-burden age groups for the number of prevalent cases and YLD counts among both genders in G20 countries remained stable at 60–89 years (Figure 2). From 1990 to 2021, the core age groups bearing the burden of prevalent cases and YLDs in both China and G20 countries showed a distinct aging trend, with the elderly emerging as a high-risk group for spinal fractures (Figure 2 and Figure 7).

**Figure 3.**
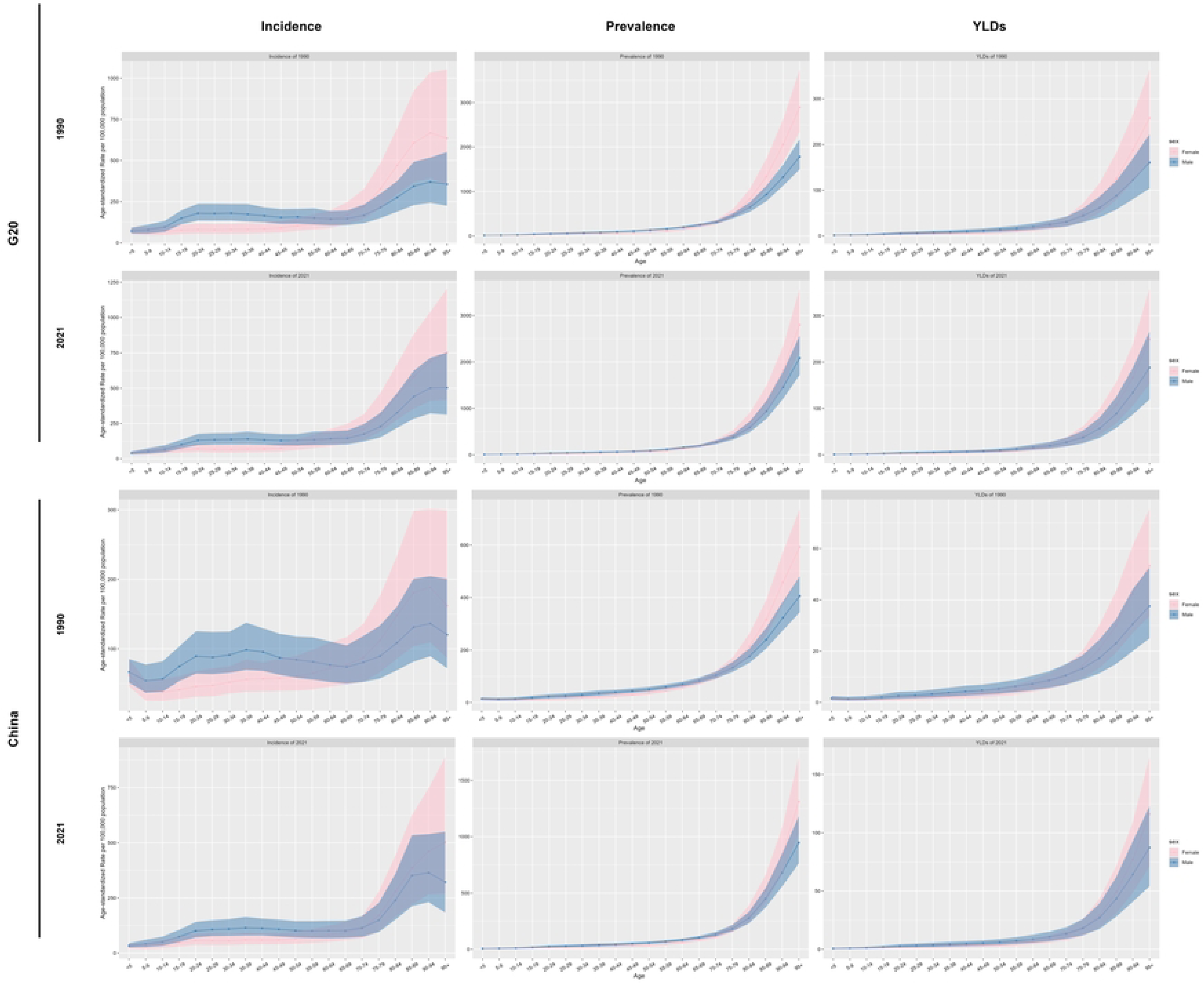
Age-sex trend line chart of disease burden of spinal fractures by age group in China and G20 countries in 1990 and 2021. (A) Age-sex trend line chart of ASIR, ASPR and ASYR of spinal fractures by age group in G20 countries in 1990 and 2021 (red for females, blue for males). (B) Age-sex trend line chart of ASIR, ASPR and ASYR of spinal fractures by age group in China in 1990 and 2021 (red for females, blue for males).

#### 3.3.3. Burden Changes Reflected by Age-Standardized Rates

From 1990 to 2021, both China and G20 countries exhibited a consistent pattern: the older the age group, the higher the age-standardized rate (Figure 6). In 1990, the burden of ASIR among Chinese males showed a bimodal distribution (35–39 years and 90–94 years), while for females, it peaked at 90–94 years with increasing age. By 2021, the burden of spinal fractures among both genders in China had surged after 70 years of age: the ASIR of males aged 70–84 years had grown rapidly, the growth rate of females aged ≥ 70 years had outpaced that of males, and the burden of males aged ≥ 94 years had decreased slightly (Figure 3). From 1990 to 2021, the high-burden stages of ASPR and ASYR for both Chinese males and females were ≥70 years, and the burden of females had grown more rapidly after this age. In G20 countries, the standardized rates of all indicators for both genders continued to rise with age, and the burden increased rapidly after 70 years (similar to the trend in China). In 2021, there was no downward trend in the burden of the elderly group (≥ 94 years), which remained at a consistently high level (Figure 3).

#### 3.3.4. Gender-Related Trends and Differences

From 1990 to 2021, gender-specific age-standardized incidence rates, prevalence rates, and YLD rates in both China and G20 countries showed phased fluctuations. In China, despite these fluctuations, the number of cases and standardized rates of incidence, prevalence, and YLDs have shown a clear overall upward trend, with particularly significant growth after 2010. In terms of gender differences, the values of the three indicators were mostly higher for males than for females (Figure 3 and Figure 8). In contrast, while the number of cases of the three types of health burdens in G20 countries had grown slowly, the standardized rates had generally shown a downward trend—reflecting a long-term improvement trend. Gender differences remained stable, with male indicators generally higher than female ones; however, after 2010, the number of prevalent cases and YLD counts among females in G20 countries had surpassed those among males (Figure 3 and Figure 8).

**Figure 3.**
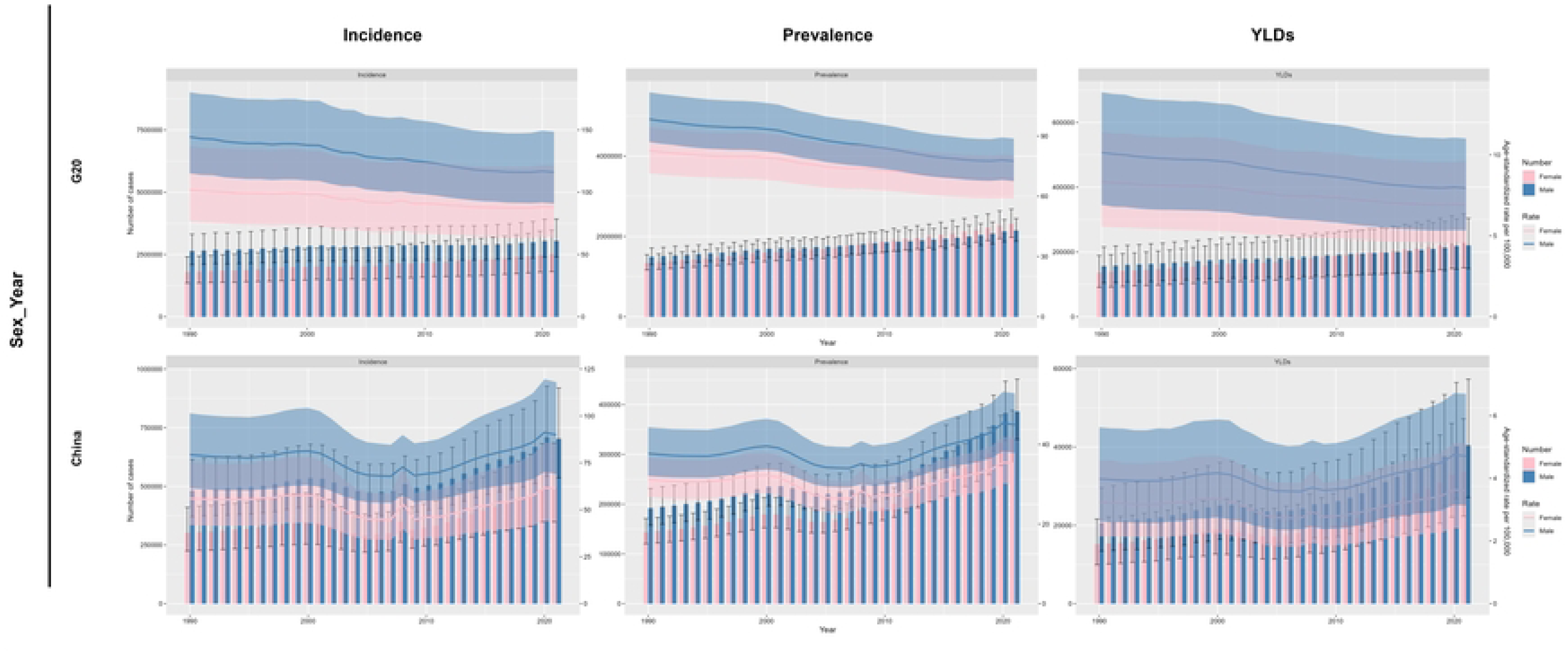
Trends in gender differences in the burden of spinal fractures in China and G20 countries from 1990 to 2021. (A) Incident cases, prevalent cases, YLDs, and age-standardized incidence rate, prevalence rate, and years lived with disability rate in G20 countries. (B) Incident cases, prevalent cases, YLDs, and age-standardized incidence rate, prevalence rate, and years lived with disability rate in China. Bar charts represent the number of cases, and line charts represent age-standardized rates (red for females, blue for males).

### 3.4. Projection of Incidence, Prevalence, and YLDs of Spinal Fractures in China and G20 countries from 2022 to 2050

It is projected that from 2022 to 2050, the ASIR of spinal fractures in Chinese males will decrease from 90.58 to 78.74, a reduction of 13.07%; the ASIR in Chinese females will decrease from 60.46 to 54.76, a reduction of 9.43%. For G20 countries, the ASIR in males will decline from 115.19 to 89.58, a decrease of 22.23%; the ASIR in females will drop from 87.71 to 75.42, a reduction of 14.01%.

The ASPR in Chinese males will increase from 45.74 to 57.13, a rise of 24.90%; the ASPR in Chinese females will decrease from 35.42 to 30.87, a reduction of 12.85%. For G20 countries, the ASPR in males will fall from 76.97 to 58.21, a decrease of 24.37%; the ASPR in females will decline from 68.35 to 55.78, a reduction of 18.39%.

The ASYR in Chinese males will increase from 4.79 to 5.93, a rise of 23.79%; the ASYR in Chinese females will decrease from 3.63 to 3.18, a reduction of 12.40%. For G20 countries, the ASYR in males will drop from 7.89 to 5.92, a decrease of 25.09%; the ASYR in females will decline from 6.86 to 5.56, an overall reduction of 18.95% (**Table 2**).

**Table 2.**
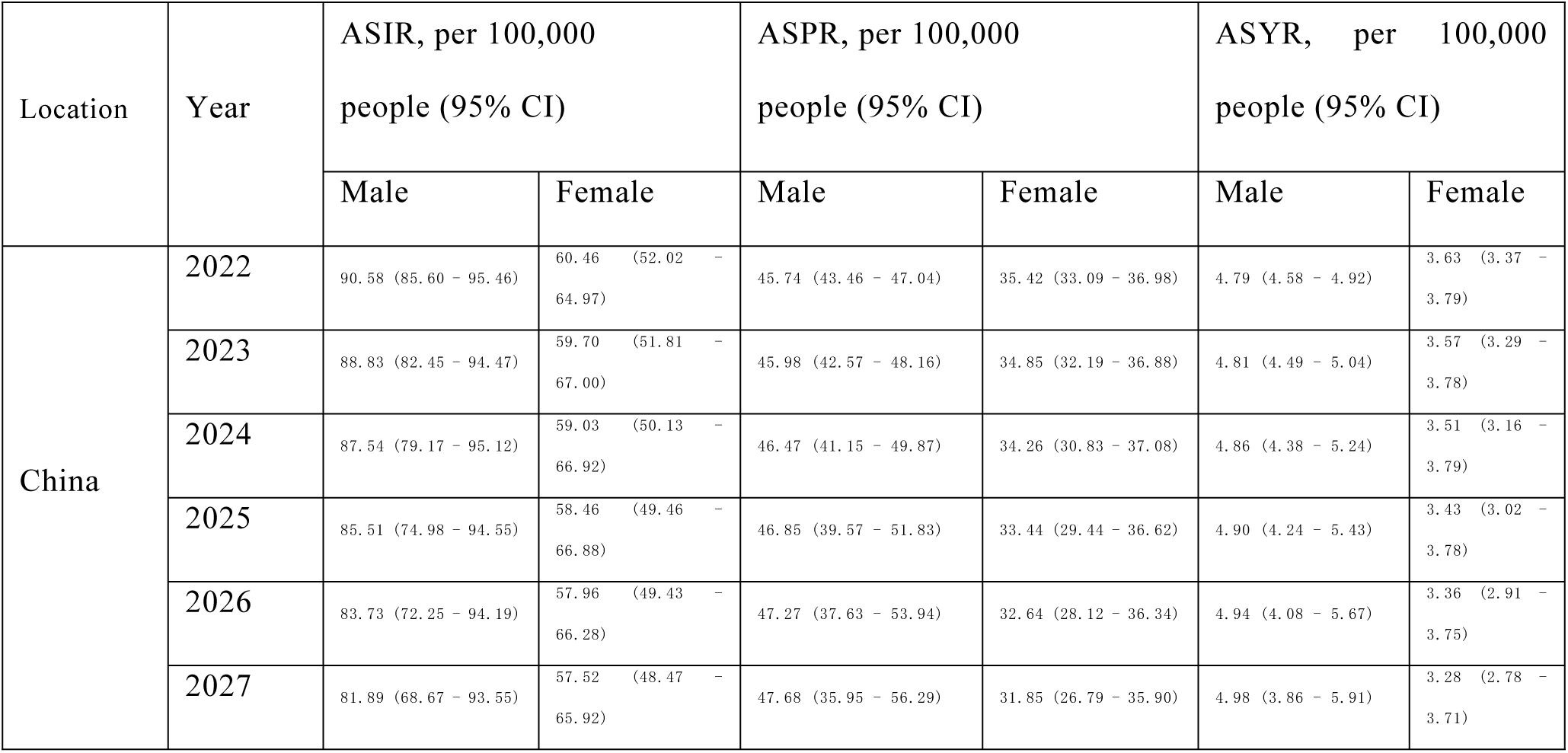
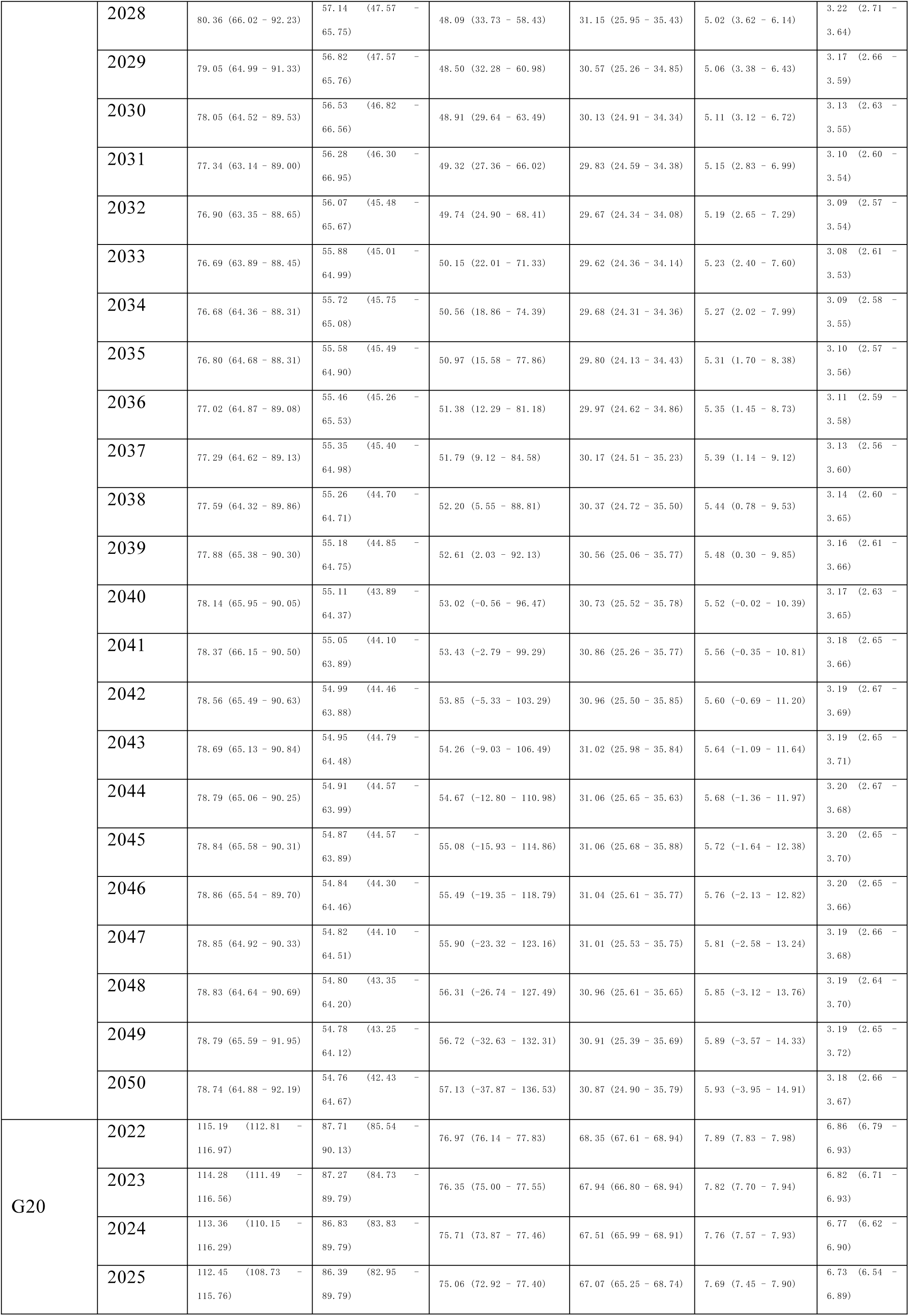
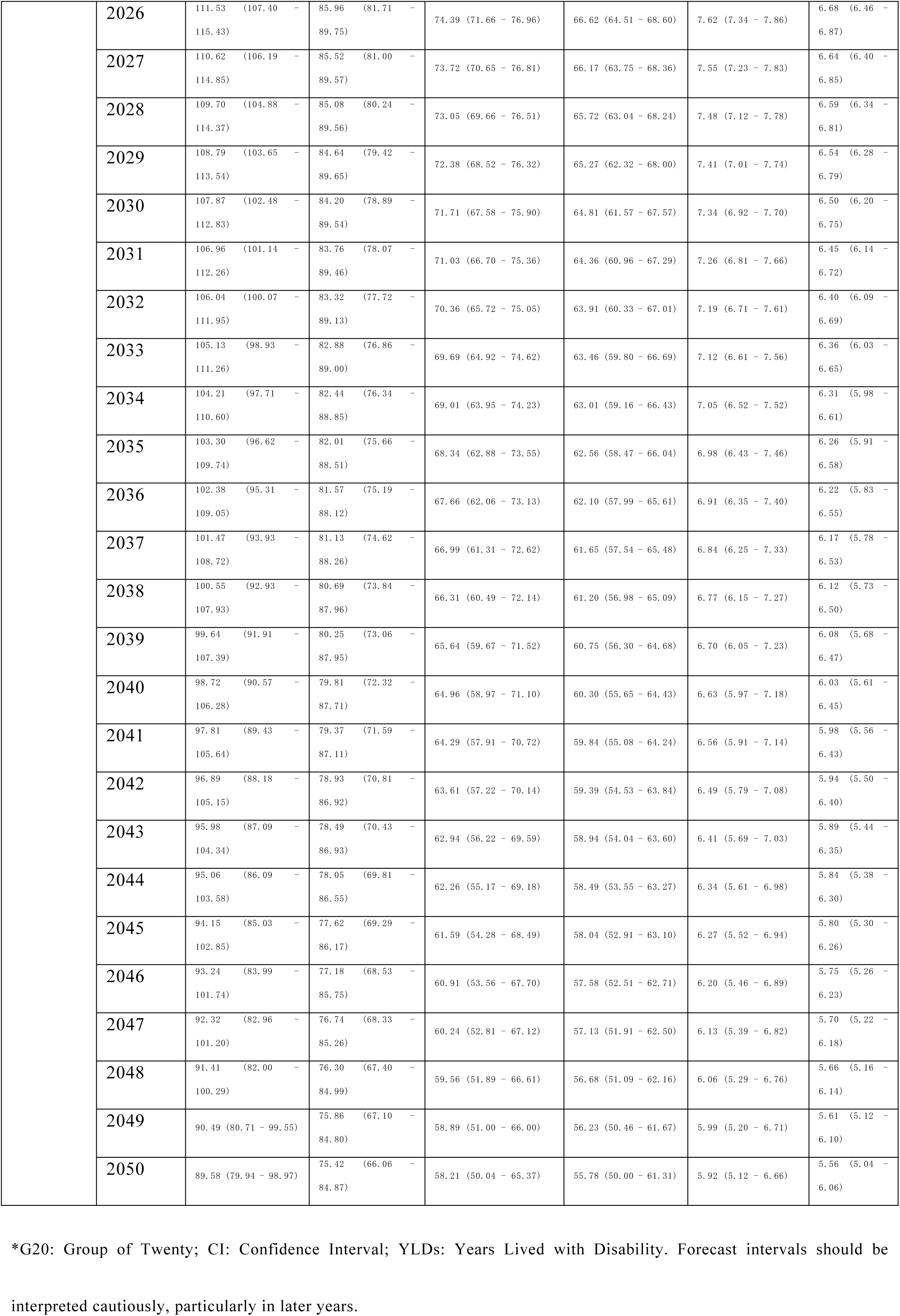
Predictions of ASIR, ASPR, and ASYR of spinal fractures in China and G20 countries from 2022 to 2050.

Overall, from 2022 to 2050, China will show significant gender differentiation: ASIR will decrease in both genders, ASPR will increase in males (by 24.90%) and decrease in females (by 12.85%), and ASYR will rise in males (by 23.79%) and fall in females (by 12.40%). In contrast, the spinal fracture-related indicators in G20 countries will exhibit the characteristic of stable decline across all indicators with consistent trends between genders (**Figure 4** and **Table 2**).

**Figure 4.**
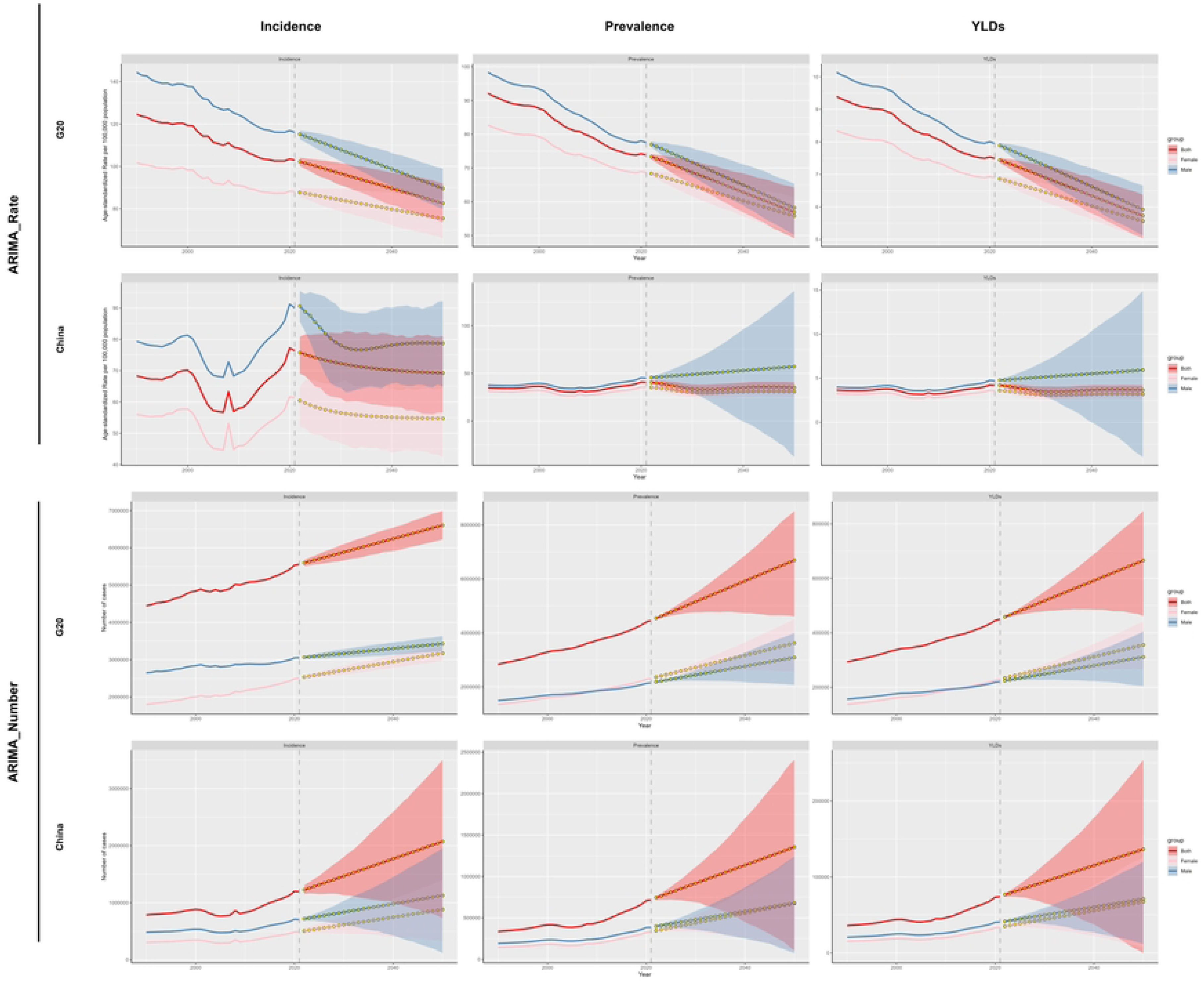
Temporal trends and 2022 – 2050 projections of ASIR, ASPR, and ASYR of spinal fractures by sex and overall in China and G20 countries from 1990 to 2021. (A) Temporal trends and projections of ASIR, ASPR, and ASYR of spinal fractures among males, females and the overall population in China. (B) Temporal trends and projections of ASIR, ASPR, and ASYR of spinal fractures among males, females and the overall population in G20 countries. Both case numbers and rates were analyzed using the ARIMA model. (red represents females, blue represents males, pink represents both sexes combined).

### 3.5. Decomposition Analysis of Spinal Fractures in China and G20 countries

In the overall G20 aggregate, epidemiological factors all acted as inhibitory factors. For incidence causes, the absolute value of G20’s population size factor exceeded that of population aging; in China, population size ranked first, while epidemiological changes and population aging had comparable values. For prevalence and YLDs causes, both G20 and China saw population aging’s value surpass population size—China’s population size value was further greater than epidemiological factors. G20’s baseline values of relevant indicators were significantly higher than China’s (Figure 5).

**Figure 5.**
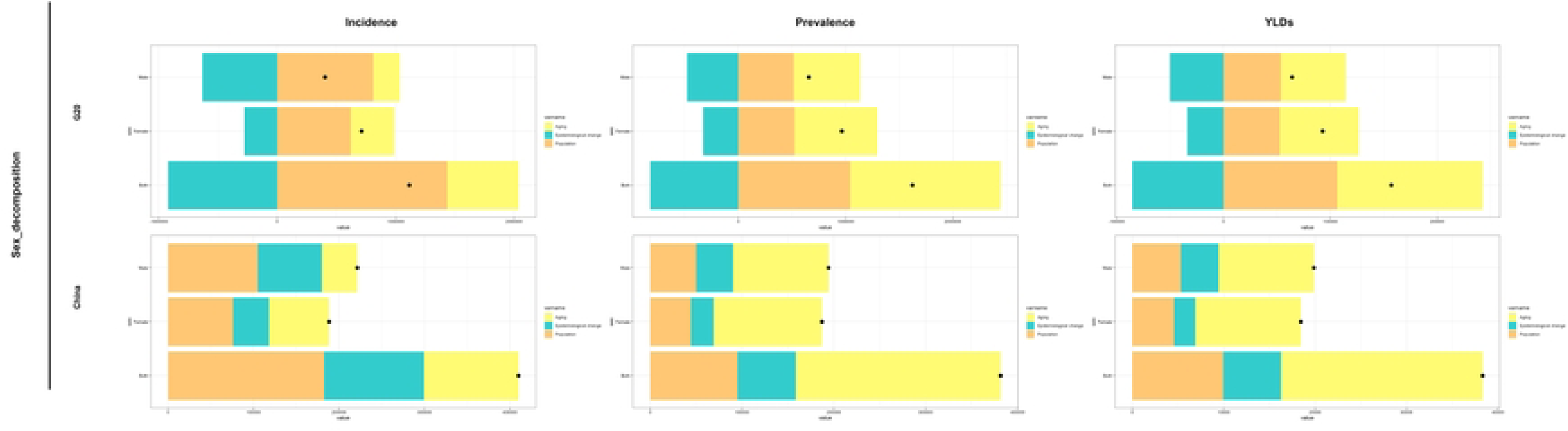
Decomposition of spinal fracture burden among males, females, and both sexes combined in China and G20 countries across incidence, prevalence, and years lived with disability (YLDs), showing the contributions of population aging, epidemiological change, and population size.

Comparative decomposition analysis between China and the overall G20 aggregate showed that for both, aging had a stronger impact on prevalence and YLDs than population factors. Comparing epidemiological values by gender, such factors affected males more than females (Figure 5).

## 4. Discussion

In this study, we systematically compared the burden of spinal fractures in China and the G20 from 1990 to 2021 and projected future trends to 2050. The main findings were that China experienced substantial increases in incident cases, prevalent cases, and YLDs over the past three decades; ASPR and ASYR increased significantly, whereas ASIR showed an overall upward tendency; in contrast, the overall G20 aggregate showed increasing absolute case numbers but declining age-standardized rates; and population aging was identified as the major driver of burden growth. These findings indicate that spinal fractures are becoming an increasingly important public health issue in China in the context of rapid demographic aging.

Spinal fractures are caused by both high-energy and low-energy trauma, with road traffic accidents and falls representing the major mechanisms in different age groups [27, 28]. In younger adults, particularly men, traumatic spinal fractures are commonly associated with motor vehicle collisions and frequently involve the cervical or thoracolumbar spine [27, 28]. Spinal cord injury is a major complication of thoracolumbar fractures and is associated with poor outcomes [28, 29]. In contrast, older adults are more susceptible to low-energy trauma-related fractures because of osteoporosis, impaired balance, and reduced physiological reserve [27]. These epidemiological and clinical characteristics help explain why the burden of spinal fractures increasingly shifts toward older populations.

An important finding of this study is that the burden of spinal fractures in China did not increase in a strictly linear manner. During 2000-2005, both ASPR and ASYR decreased significantly, whereas all three age-standardized indicators increased after 2010. These temporal fluctuations may reflect changes in injury exposure, healthcare access, diagnostic practices, and broader social or policy contexts; however, such explanations cannot be directly tested in the present ecological analysis. Nevertheless, the concurrent increase in ASIR, ASPR, and ASYR in recent years suggests that the burden of spinal fractures in China has increased and has diverged from the overall downward trend in age-standardized rates observed in the overall G20 aggregate.

Our age-specific analyses further showed that the burden of prevalent cases and YLDs in China was increasingly concentrated in older adults of both sexes. This pattern is likely related to rapid population aging, improved survival after fracture, and the accumulation of chronic disability over time. Although the overall G20 aggregate also showed a concentration of burden in older populations, the age distribution in these countries appeared relatively more stable during the study period. Notably, age standardization did not fundamentally alter the identification of older adults as the core burden-bearing population in China, suggesting that the increasing burden reflects not only demographic change but also persistently high age-specific risks in later life. Age-related bone loss, sarcopenia, falls, multimorbidity, and improved case detection are all likely contributors to this pattern [27, 28].

We also observed substantial sex differences in spinal fracture burden. In both China and G20 countries, males generally had higher age-standardized incidence, prevalence, and YLD rates at younger ages, whereas in older age groups, especially after menopause, the burden among females increased more rapidly. This pattern is biologically plausible, as postmenopausal estrogen decline accelerates bone loss and substantially increases the risk of osteoporotic fracture. Previous studies have also shown that low bone mineral density is a major determinant of vertebral fracture risk and that osteoporosis is highly prevalent among older women [30]. Beyond demographic aging, epidemiological risk factors such as low bone mineral density, obesity, falls, polypharmacy, low physical activity, weak muscle strength, smoking, low educational level, and hypoestrogenemia may further increase fracture risk [7, 31–33].

The etiology of spinal fractures varies substantially across populations. In young and middle-aged adults, especially men, high-energy trauma such as falls from height and traffic accidents remains the dominant cause [28], whereas the elderly are more often affected by osteoporotic vertebral compression fractures related to reduced bone strength and increased skeletal fragility [34, 35]. Pediatric spinal fractures show yet another pattern, with trauma-related mechanisms still predominating [36]. This age-stratified etiological profile suggests that prevention strategies should be population-specific: trauma prevention should be emphasized in children and younger adults, whereas osteoporosis screening, fall prevention, and bone health management should be prioritized in older populations. It should be noted that the GBD 2021 database aggregates osteoporotic, traumatic, and pathological fractures into a single category, which limits the ability to analyze subtype-specific trends. Despite this limitation, reporting the combined burden provides a comprehensive population-level assessment and allows meaningful comparison between China and the G20 aggregate. Age-stratified analyses include pediatric populations according to the GBD methodology, which may introduce heterogeneity in fracture risk profiles across age groups.

Screening and intervention for osteoporosis are central to reducing the future burden of spinal fractures. Current evidence supports routine risk assessment and screening in high-risk populations, particularly older women, while recognition of osteoporosis in men also remains important [37–40]. Preventive strategies should include adequate calcium and vitamin D intake, balance and strength training, and pharmacological treatment when indicated, such as bisphosphonates or denosumab [41]. From a clinical perspective, management should be individualized according to fracture type and patient characteristics. For acute vertebral compression fractures, conservative treatment and long-term osteoporosis management remain important, while vertebroplasty may be considered in selected cases [42, 43]. In patients with multiple non-contiguous spinal fractures, comprehensive spinal imaging and stabilization of injured segments are essential [44]. In children, clinicians should remain alert to spinal cord injury without radiographic abnormality and pursue additional imaging when clinically indicated [45].

The economic consequences of spinal fractures are also substantial. In China, direct medical costs of vertebral fractures increased markedly from 2013 to 2017, underscoring the growing pressure placed on individuals, families, and the healthcare system [6]. Our projections suggest that although China’ s ASIR may gradually decline, ASPR and ASYR may remain elevated, particularly among males. This pattern indicates that even if the rate of new occurrence slows, the long-term burden of prevalent disease and disability may continue to accumulate. Such a trend may reflect improved survival after fracture, persistent rehabilitation needs, under-recognition of osteoporosis in men, and gender-related differences in health-seeking behavior [39, 46]. By contrast, the overall G20 aggregate is projected to show a more consistent decline in age-standardized burden. These projection results should, however, be interpreted cautiously because long-term forecasting inevitably involves uncertainty.

This study has several limitations. First, as with all GBD-based analyses, the estimates depend on the quality and completeness of the underlying data sources; underreporting and misclassification may persist, especially in settings with limited primary healthcare or surveillance capacity [47]. Second, the GBD database does not distinguish spinal fractures by specific etiology, such as osteoporotic, traumatic, or pathological fractures, which limits subtype-specific interpretation. Third, this was an ecological study based on population-level estimates; therefore, causal inferences regarding temporal changes and cross-national variation should be made with caution. Fourth, substantial regional heterogeneity exists within China in healthcare resources, diagnostic capacity, and data quality, but these differences could not be fully evaluated in the current analysis. Furthermore, the GBD dataset does not provide detailed diagnostic criteria for spinal fractures. For osteoporotic vertebral fractures, cases may be identified based on radiographs or clinical symptoms, which may or may not lead to imaging. This variability could contribute to differences in reported incidence and prevalence, limiting the interpretability of trends across countries and age groups.

Moreover, because the pooled G20 estimates include China, the comparison between China and the G20 aggregate should be interpreted as a descriptive comparison at the group level rather than fully independent between-group contrasts. This approach allows contextualizing China’ s burden within the overall G20 trends. “ Additionally, the heterogeneity of fracture types and the inclusion of all age groups, including children, should be considered when interpreting the results, particularly for subtype-specific or age-specific trends.

Overall, our findings indicate that the burden of spinal fractures in China has increased substantially over the past three decades and differs from the declining age-standardized trends observed in the overall G20 aggregate. Population aging appears to be the dominant driver of this increase, while older adults and sex-specific high-risk groups represent key targets for intervention. Strengthening osteoporosis screening, fall prevention, rehabilitation, and long-term care capacity will be essential for reducing the future burden of spinal fractures in China.

## 5. Conclusion

From 1990 to 2021, the burden of spinal fractures in China increased substantially, with marked rises in incident cases, prevalent cases, and years lived with disability (YLDs). The age-standardized prevalence rate (ASPR) and age-standardized YLD rate (ASYR) showed significant upward trends, whereas the age-standardized incidence rate (ASIR) showed an overall upward tendency. In contrast, the overall G20 aggregate showed declining age-standardized rates despite increases in absolute case numbers. Population aging was the principal driver of burden growth, particularly for prevalence and disability burden. Older adults remained the main high-risk population, and clear age- and sex-specific differences were observed. Projections to 2050 suggest that although some indicators may decline, China may continue to face substantial challenges in the long-term prevention and management of spinal fractures under rapid demographic aging. Strengthening osteoporosis screening, fall prevention, rehabilitation, and age-specific intervention strategies may help reduce the future burden of spinal fractures in China.

## Data Availability

No new data were generated in this study. All data used are publicly available from the Global Burden of Disease (GBD) 2021 study. Data can be accessed through the Global Health Data Exchange (GHDx) at: https://ghdx.healthdata.org/gbd-2021.

## Abbreviations

YLDs: years lived with disability;
GBD: Global Burden of Disease;
ASRs: age-standardized rates;
ASIR: age-standardized incidence rate;
ASPR: age-standardized prevalence rate;
ASYR: age-standardized YLD rate;
EAPC: estimated annual percentage change;
CI: confidence interval;
APC: age–period–cohort;
ARIMA: autoregressive integrated moving average.

## Appendix figure

**Figure 6.**
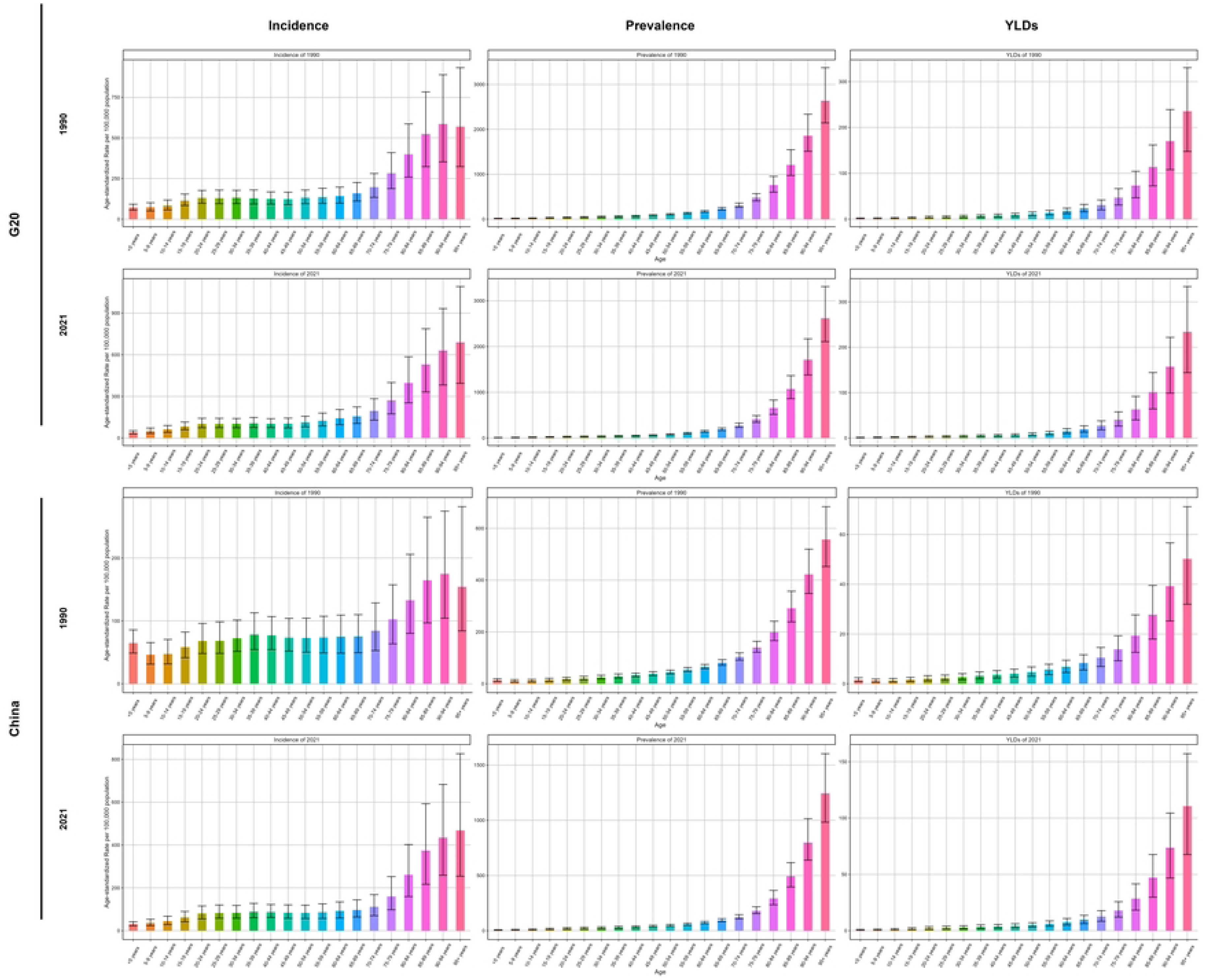
ASIR, ASPR, and ASYR of spinal fractures by age group for both sexes combined in China and G20 countries in 1990 and 2021. (A) ASIR, ASPR, and ASYR by age group in G20 countries in 1990 and 2021. (B) ASIR, ASPR, and ASYR by age group in China in 1990 and 2021.

**Figure 7.**
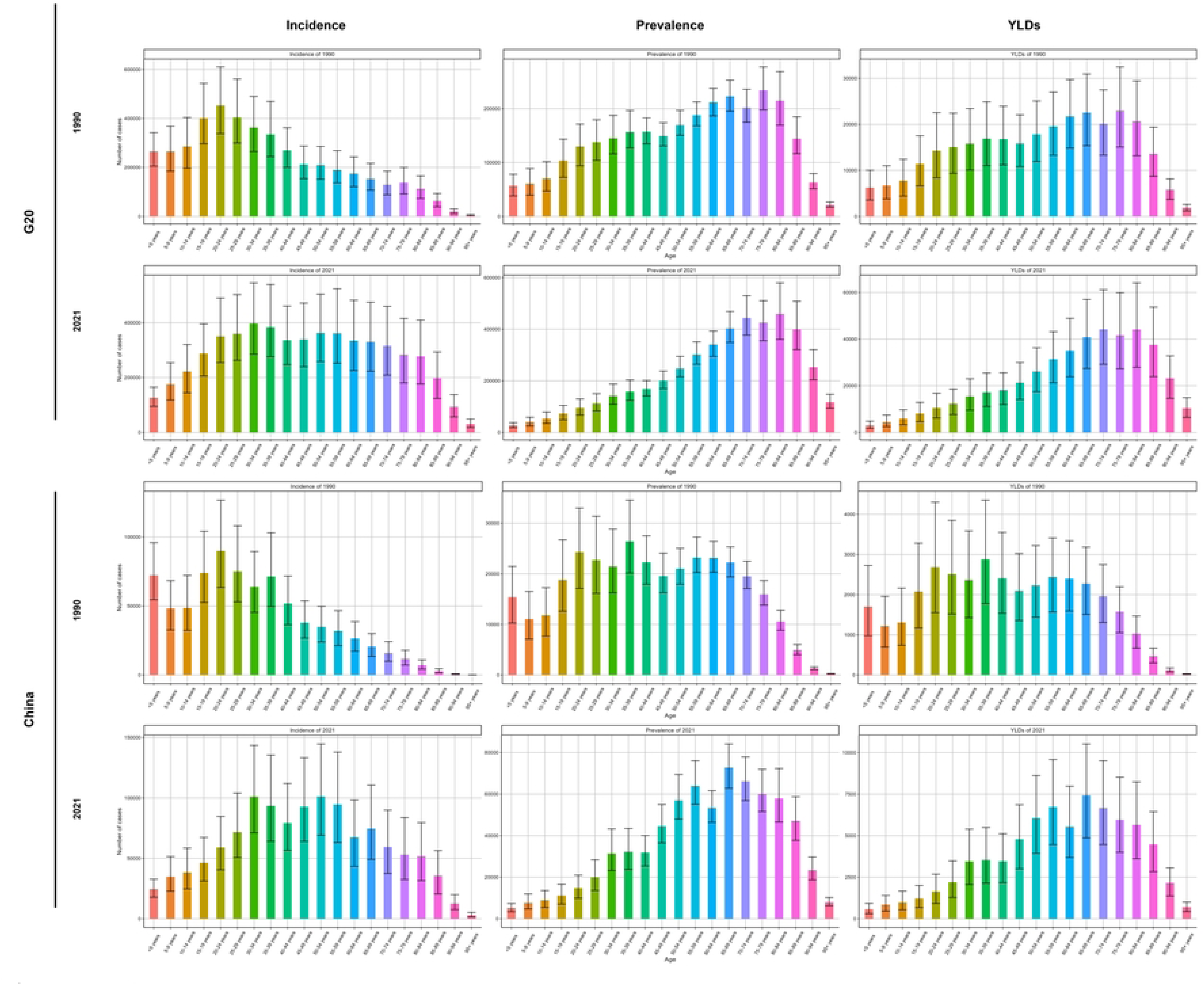
Numbers of incident cases, prevalent cases, and YLDs of spinal fractures by age group for both sexes combined in China and G20 countries in 1990 and 2021. (A) Number of incident cases, prevalent cases, and YLD counts by age group in G20 countries in 1990 and 2021. (B) Number of incident cases, prevalent cases, and YLD counts by age group in China in 1990 and 2021.

**Figure 8.**
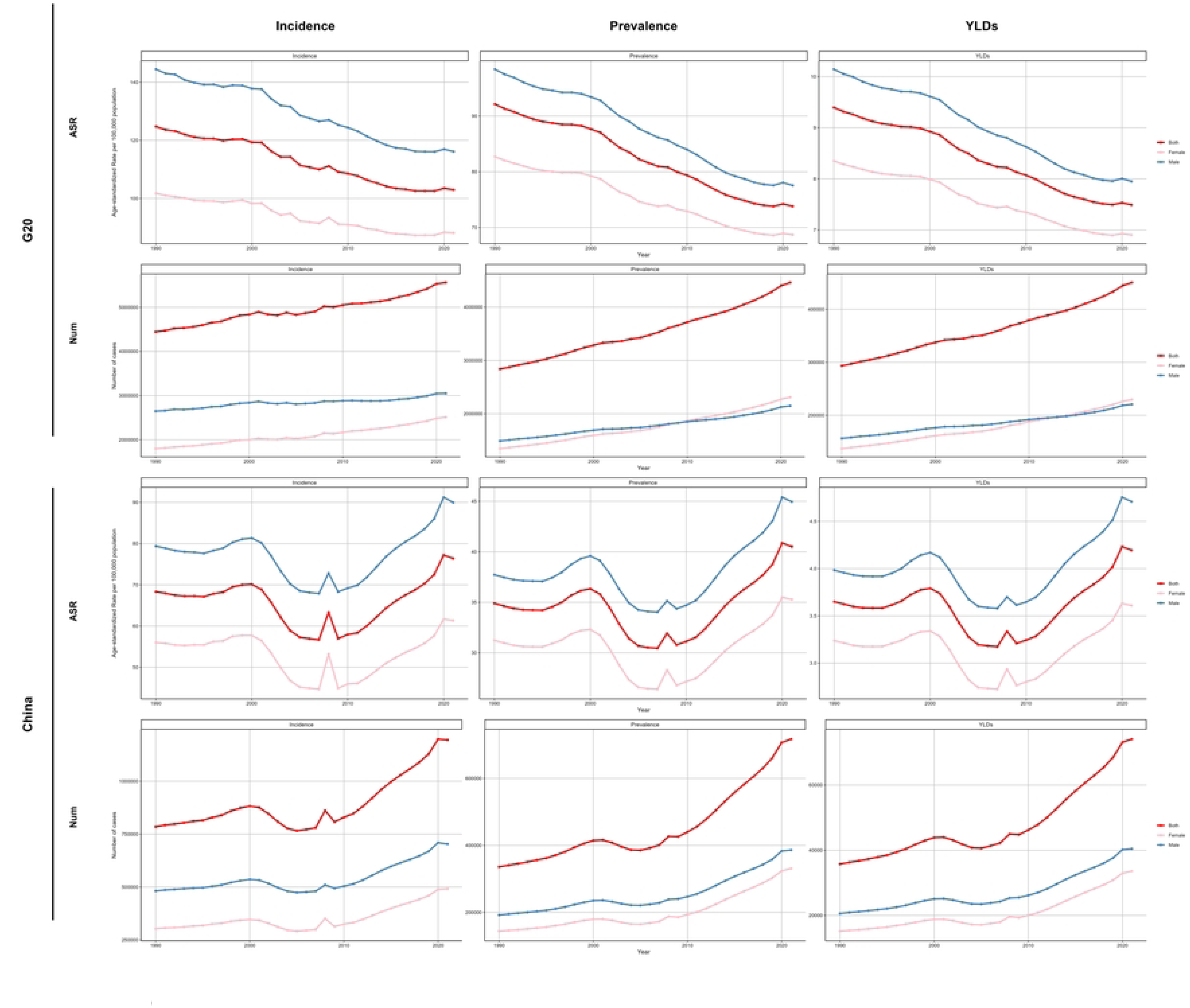
Comparison of ASIR, ASPR, and ASYR of spinal fractures by age group and sex (males, females, and both sexes combined) between China and G20 countries from 1990 to 2021. (A) ASIR, ASPR, and ASYR of spinal fractures by age group in G20 countries from 1990 to 2021. (B) ASIR, ASPR, and ASYR of spinal fractures by age group in China from 1990 to 2021 (red represents females, blue represents males, pink represents both sexes combined).

## Availability of Data and Materials

The data used in this study are publicly available on the website of the Institute for Health Metrics and Evaluation (IHME) (https://ghdx.healthdata.org/gbd-2021).

## Consent for Publication

Not applicable.

## Informed Consent

This study uses de-identified data from the GBD 2021 public database. The database complies with ethical standards, with informed consent obtained from all original participants. As a secondary analysis of publicly available data without direct human subject involvement, no additional informed consent is required.

## Clinical Trial Registration

Clinical trial number: Not applicable.

## Funding

No funding was received for the research, writing, and/or publication of this article.

## Author Declarations

All claims expressed in this article are solely those of the authors and do not necessarily represent those of their affiliated organizations, or those of the publisher, the editors, and the reviewers. Any product that may be evaluated in this article, or claim that may be made by its manufacturer, is not guaranteed or endorsed by the publisher.

## Acknowledgments

We thank the Institute for Health Metrics and Evaluation (IHME) for providing open access.

## Conflicts of Interest

Shi-Jie Zeng^1^, Jia-Lan Chen^2^, Zhao-Feng Lin^3^, Jia-Qi Zhang^1^, and Li-Xin Zhu^1,^* declare that they have no commercial or financial relationships that could be construed as potential conflicts of interest.

## Notes

### Competing Interest Statement

The authors have declared no competing interest.

### Funding Statement

The author(s) received no specific funding for this work.

### Author Declarations

This study used publicly available, de-identified data from the Global Burden of Disease (GBD) 2021. Ethical approval was not required, as the analysis involved no human participants, patient data, or identifiable information.

